# A Full-Scale Agent-Based Model of Lombardy COVID-19 Dynamics to Explore Social Networks Connectivity and Vaccine Impact on Epidemic

**DOI:** 10.1101/2020.09.13.20193599

**Authors:** Giuseppe Giacopelli

## Abstract

September 13, 2020

COVID-19 outbreak is an awful event. However it gives to the scientists the possibility to test theories about epidemic. The aim of this contribution is to propose a individual-based model of Lombardy COVID-19 outbreak at full-scale, where full-scale means that will be simulated all the 10 millions inhabitant population of Lombardy person by person, in a commercial computer. All this to test the impact of our daily actions in epidemic, investigate social networks connectivity and in the end have an insight on the impact of an hypothetical vaccine.

## 1 Introduction

The first alarm from COVID-19 has arrived from China [1], but one of the most dramatic outbreak was in Italy at the end of January [2]. This epidemic was the setting of a change in risk management: the use of mathematical modeling [3]. In this direction must be mentioned the paper of [4], where is explored the COVID-19 outbreak in agent-based model of about 750 thousands inhabitants of the city of Urmia, with the movement of agents approximated by places. On the same direction is the contribution [5], where is taken in account a model of transmission with a sub-sampled population of 9 thousands people living in Daegu. But there are also small-scale models as [6]. Must be also mentioned the model developed by University of Palermo [7] based on the paper [8].

The aim is to introduce a qualitative full-scale agent-based model able to reproduce the Lombardy COVID-19 dynamics, modeling its outbreak and descent, including as much real data as possible. Being Lombardy population about 10.06 millions makes this model very large scale in comparison with previous contributions. In second instance will be taken in account several alternative scenarios, in order to weight the events of those days. Then the model of social interaction used in epidemiological simulations, will be explored in terms of graph theory [9], in order to analyze it as a social network [10]. In conclusion will be drawn several conclusions about the impact of our habits in COVID-19 outbreak.

## 2 Methods

### 2.1 Model structure

The key idea of the model is to create a three layers model, as can be seen in figure 1. The first layer is an agent based particle model of Lombardy. Every agent is an inhabitant of the region, making this model a full-scale model of the Italian region. Then we have 10.06 millions agents that move with a random walk [11]. This little detail is part of the novelty of the contribution, because (at the best of author’s knowledge) it is the first try to simulate a so large population individual by individual for this purpose.

**Figure 1:**
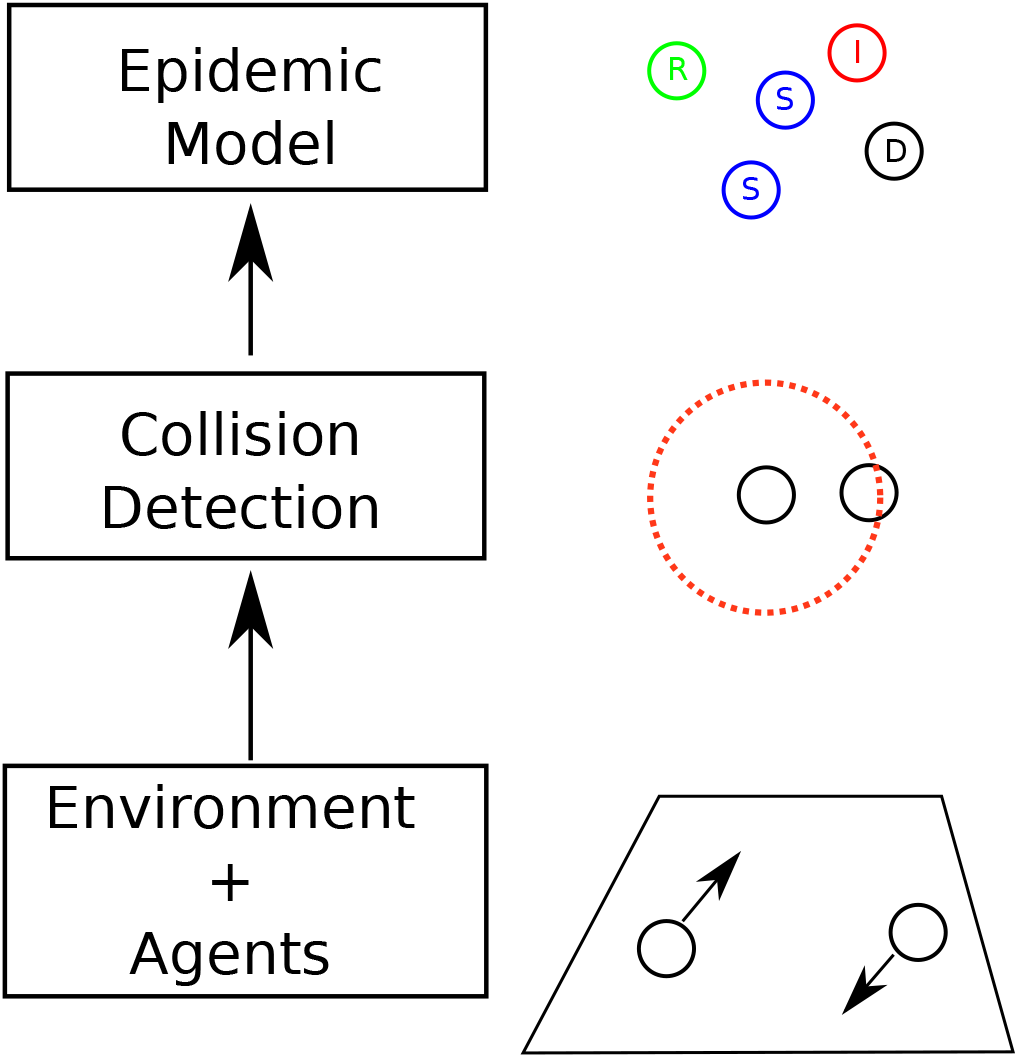
Three layers structure of the model.

On the agents model is built a collision detection algorithm, able to detect if two particles have distance less than a fixed value. However the large-scale reached by the model will require an ad hoc algorithm for this purpose, that will be a challenging problem.

On the collision detection model is built the epidemic model, that is a SIRD (Susceptible-Infected-Recovered-Dead) model [12]. The model has been filled with many experimental findings on COVID-19 and some fitting parameters are fitted on experimental findings. In the whole analysis have been used as much open access contributions as possible, moreover the whole project is fully open source at the site [13].

### 2.2 Agents model

The model simulates the behavior of each inhabitant of Lombardy using the approximation of Random walk [11]. Then every particle moves with a random vector at every step. In particular the model runs at 6 Frames per days, that is a good frame rate considering that the scale time for Epidemic phenomena is usually months. The approach of random walk can appear unrealistic, but this hypothesis ha shown to be appropriate to model very large scale systems, like in gas thermodynamics [14]. In addition to the random walk, has been added a weak velocity field with a dependence of 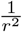, where *r* is the distance between two particles, like in gravitational field, in order to aggregate particles. The drift speed of the particles is constant and selected with a Weibull distribution [15] with scale parameter 6 and shape parameter 1.5. The particles speeds has been adjusted through a multiplicative constant in order to make the average path length of a particle in a day about 43 Km, as suggested in the report [16].

### 2.3 Environment model

The setting of the simulation is Lombardy, then the environment model will be a closed 2D box with boundary shape following Lombardy border. In order to keep particles inside the region, has been introduced a bouncing condition at the border, so that a particle that tries to cross the border bounces backward. This condition is very popular in gas thermodynamics [11]. The initial conditions of particles in terms of position are distributed following the actual density of population of Lombardy, extracted from the image [17] with techniques of image analysis and then must be intended as an approximation of the real data.

### 2.4 Collision detection

Starting from the assumption that the algorithm has been designed to run on a commercial computer in parallel (the one used has a AMD 3900X 12-cores and 64 GB of RAM) and in reasonable time (about 20 minutes of calculation for 14 days of simulation), the collision detection algorithm had a central role in the implementation of the algorithm. Because to find all the points with distance less than a constant in a set of *N* points, in general has complexity order of *N*^2^. In our model *N* ≈ 10^7^, then the complexity is order of 10^14^, that’s an incredible number!

Then the idea is to subdivide the Lombardy with a grid with thick about 20 Km and in every cell of the grid apply the collision detection, all this in parallel through cells. This multi-scale processing has lead to a speed up of the code, reducing at the same time the RAM used simultaneously in computation, making possible a simulation with 10 millions particles at the same time. However this approach neglects all the connections across the borders of the cells, the connections detected fraction (if the detection distance is about 1 Km) has been estimated computationally on a small scale experiment to be about 97.8% [13], that is a good percentage in a qualitative model. However this percentage can be increased using a multiple grids detection scheme, but it is beyond the aims of this contribution.

### 2.5 Epidemic model

The Epidemic model is a SIRD (Susceptible-Infected-Recovered-Dead) model [3]. Most of the models available up to now are called population models [20]. A population model is a model where every node is modeled by a set of differential equations and it models a sub-population of a region. The number of people modeled by a single node can range from hundreds to millions. In our model, every node is a single person. So the model is not an ODE (Ordinary Differential Equations) model, but a stochastic agent-based model. Every node has 4 states:

1. Susceptible: it hasn’t already contracted the disease. It can be become Infected with a probability *p_I_* for each contact with an Infected;
2. Infected: it is infected and then it can infect Susceptible nodes. After *E* days will change its state to Recovered or to Dead, in particular it has probability *p_D_* to die and 1 − *p_D_* to be recovered;
3. Recovered: it has been healed from the disease and it can’t contract it either infect Susceptible nodes any more;
4. Dead: the node has died, so it can’t infect any more.

## 3 Results

### 3.1 Lombardy Outbreak

First of all I want to point out that all the simulations are available in .avi format at the link [13], as well the MATLAB code to reproduce them. Then the first scenario is the Lombardy outbreak of March 2020 [2]. Our simulation starts 29 February 2020 and terminates 14 March 2020. The main realistic parameters are 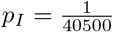 (extracted from the report [21]) and *p_D_* = 0.3 (estimated from Worldometer [22] that has been cited also in [23]).

The fitted parameters are the collision radius of 1 Km (that can appear enormous comparing to the 1 m distance suggested by WHO [24], but must be taken in account that there are 6 frames per day, then 1 Km is the radius of interaction of a person that stays in the same place for 4 hours) and the days of disease *E* = 7 ([25] suggests *E* < 10).

The results of simulations can be seen in figure 2. The model is able to explain the experimental data until about 9 March 2020. What happened after this date belongs to history. Indeed 9 March was the day of the DPCM “Misure urgenti in materia di contenimento e gestione dell’emergenza epidemiologica da COVID-19” [19], a law in order to contain the COVID-19 outbreak. This law was the beginning of the Italian lock down. In this discrepancy between the data and the model, can be seen all the efforts of Italian people to block the Coronavirus. This simulation also warns us on what would have could happened.

**Figure 2:**
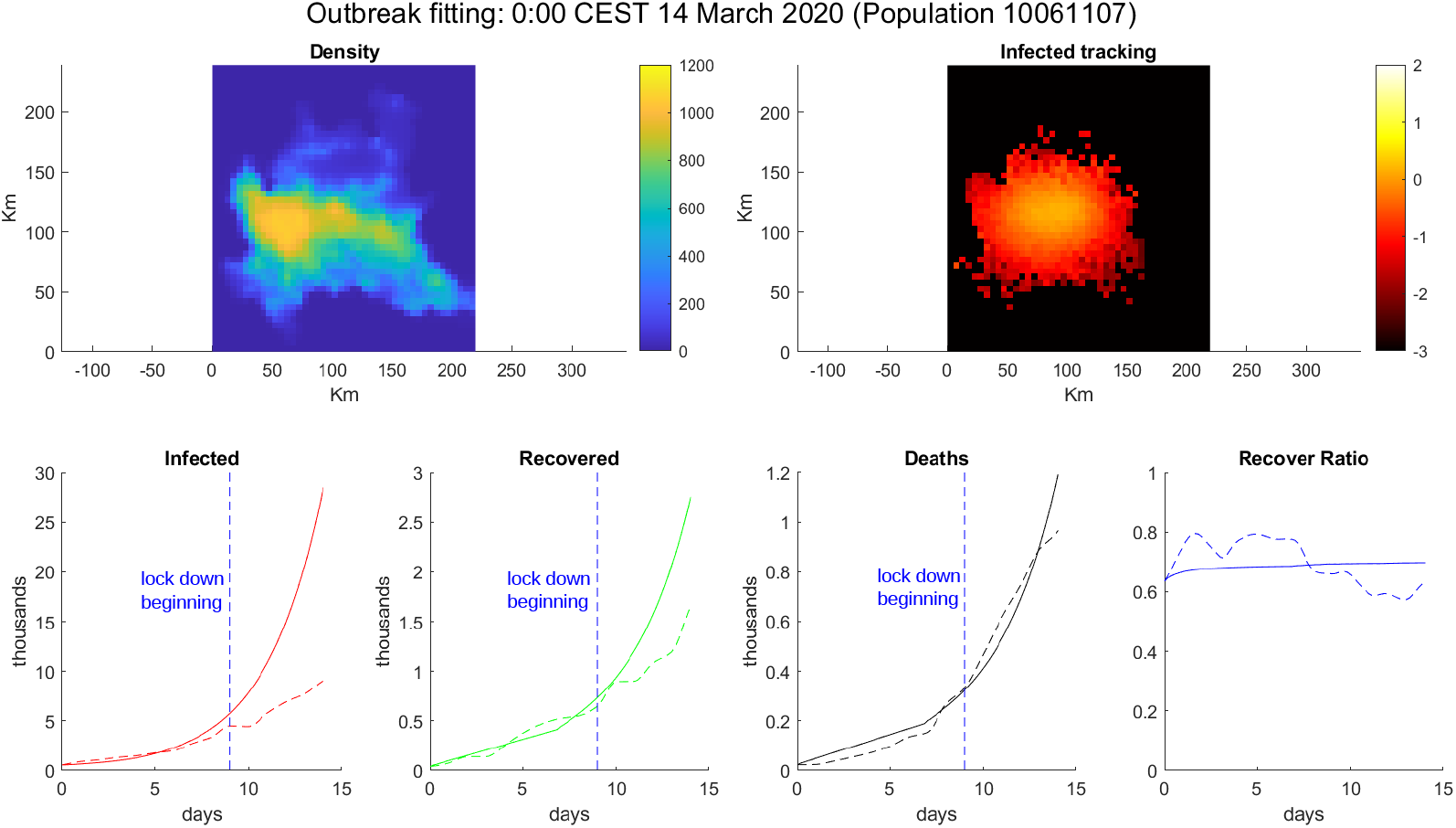
Simulation of COVID-19 Outbreak. (Top-Left) Density of Population. (Top-Right) log_10_ of infected percentage per cell. (Bottom) From left to right: Infected number, Recovered number, Deaths number and Recover ratio 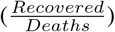. Solid line is model simulation, dotted line is extracted data by [18] for Lombardy and vertical dotted blue line is the date of DPCM 9 March [19].

**Figure 3:**
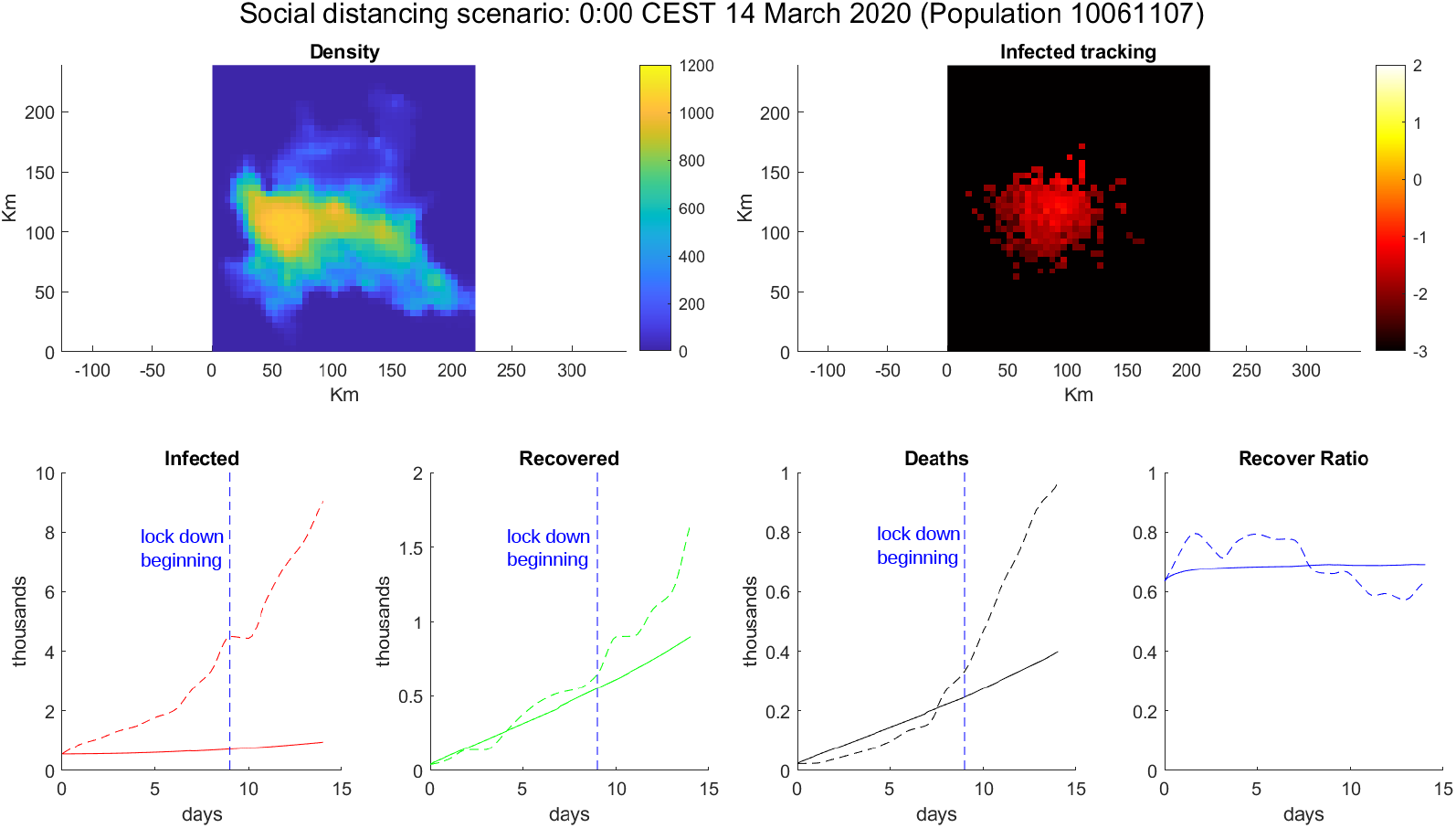
Simulation with social distancing. (Top-Left) Density of Population. (Top-Right) log_10_ of infected percentage per cell. (Bottom) From left to right: Infected number, Recovered number, Deaths number and Recover ratio 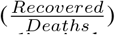. Solid line is model simulation, dotted line is extracted data by [18] for Lombardy and vertical dotted blue line is the date of DPCM 9 March [19].

### 3.2 Our habits make the difference

This second scenario is inspired from the paper [26]. In this paper the authors show that keeping a 2 meters distance between people halves the risk of contracting COVID-19. Then the aim of this simulation is to simulate this kind of social distancing by halving *p_I_* in the model. The results, in figure 3, is that the COVID-19 (in this particular scenario) isn’t enough contagious to spread as in the experimental data. This simple simulation shows the striking role that we have in fighting this disease, indeed a simple action, like to keep a social distance, can make the difference between a virus under control and an epidemic. This simple fact has already been observed in experimental finding about Germany [27], where a synthetic method was used to estimate the contagion without the use of masks. On the same path has been performed a simulation of lock down, reducing the daily average kilometers done by a node from 43 to 5 and reducing the interaction distance from 1 Km to 100 m. The results of this simulation can be seen in figure 4. These simple assumptions were enough to defeat the virus.

**Figure 4:**
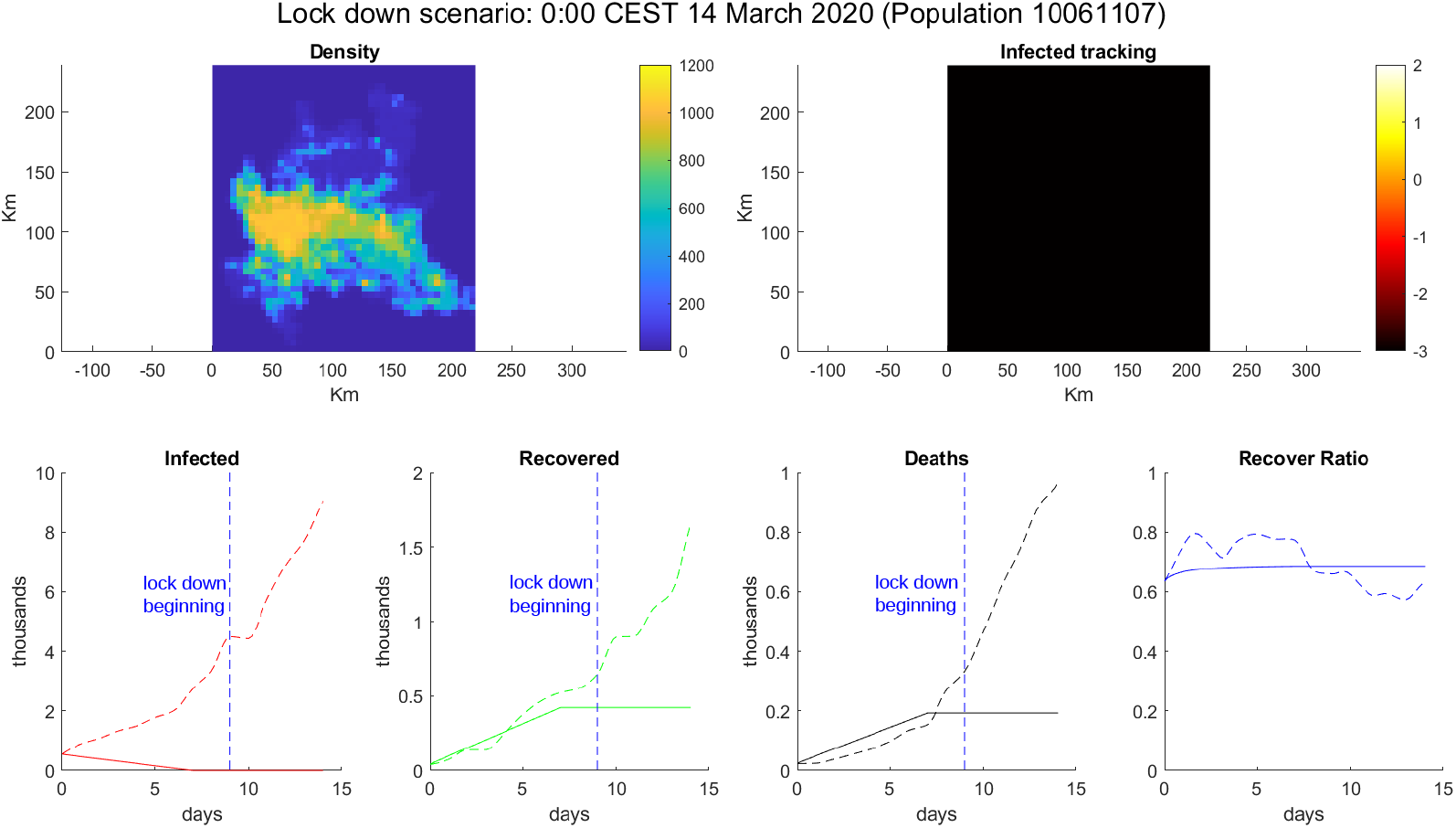
Simulation of lock down. (Top-Left) Density of Population. (Top-Right) log_10_ of infected percentage per cell. (Bottom) From left to right: Infected number, Recovered number, Deaths number and Recover ratio 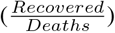. Solid line is model simulation, dotted line is extracted data by [18] for Lombardy and vertical dotted blue line is the date of DPCM 9 March [19].

### 3.3 Network Topology

The impact of topology in an epidemic model is an hot topic [28] in the debate about social networks. For this reasons has been performed a test, one thousands particle has been chosen and then tracked in all the simulation to find the total number of connection with the whole population. In graph theory, the number of connection of a node is called degree [9]. With this test has been determined the degree distribution and the daily degree distribution (average degree per day) of this small group of people varying in time. However here will be presented just the final result, for the full simulation refer to [13]. The first scenario is the COVID-19 Outbreak scenario, figure 5. Can be seen that the distribution has an evident left tail (in contrast with the right tail of BA models [10]). Probably this fact emerges from the simulation time of 14 days (in contrast with the social networks, that usually takes years to be built). It is also interesting the scenario of lock down. In this scenario we have a fall of the connectivity from thousands average connections per day to order of hundred. This results shows the importance of the policy of lock down in COVID-19 containment.

**Figure 5:**
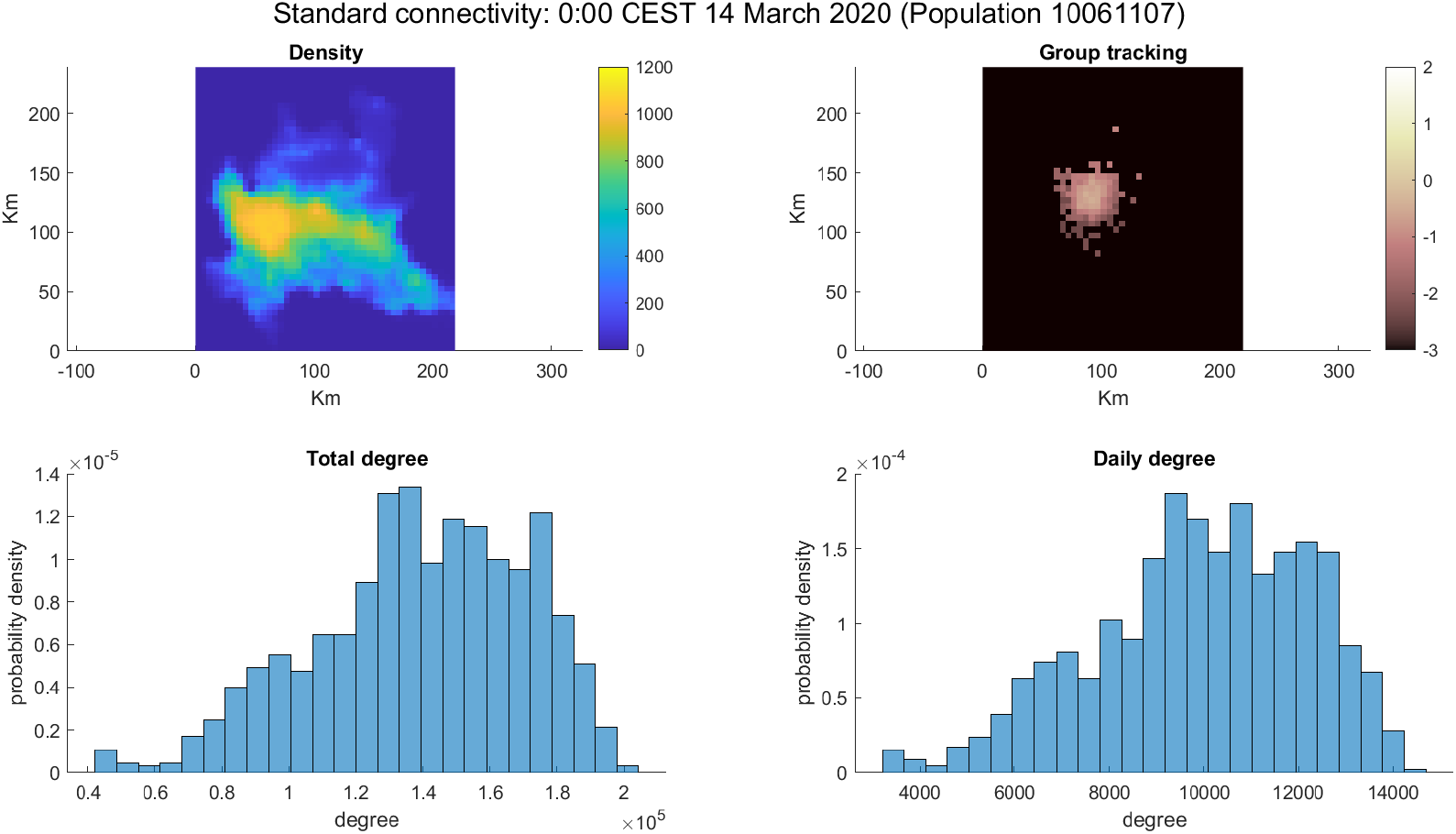
COVID-19 Outbreak simulation connectivity. (Top-Left) Density of Population. (Top-Right) log_10_ of group percentage per cell. (Bottom-left) Degree distribution of small group. (Bottom-Right) Daily degree distribution of small group.

**Figure 6:**
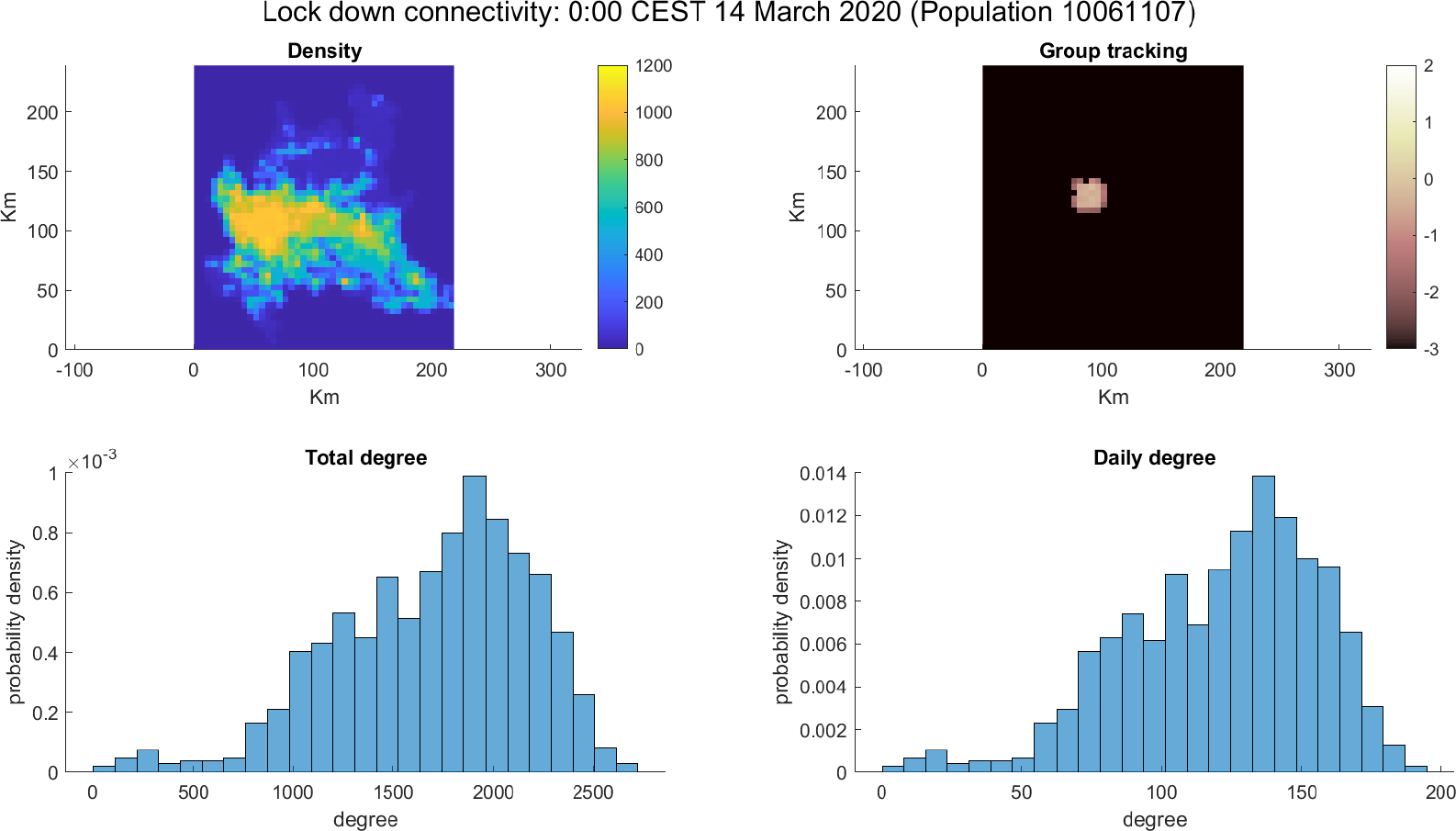
Lock down simulation connectivity. (Top-Left) Density of Population. (Top-Right) log_10_ of group percentage per cell. (Bottom-left) Degree distribution of small group. (Bottom-Right) Daily degree distribution of small group.

### 3.4 The descent

In this scenario will be taken in account the period between 31 May 2020 and 14 June 2020. In this period Italy is concluding its lock down and the active cases number is decreasing. For this simulation the kilometers per day has been arbitrary set to 15, because of the lack of additional information about mobility during this period. The probability of contagion has been halved to take in account the social distancing. The radius of interaction and the duration of the disease has been tuned to reproduce experimental data. In particular the value for radius of interaction is 300 m and as disease duration has been taken *E* = 35, so 5 weeks. This value (so higher in comparison to the one of the outbreak) could be motivated by a clinical protocol more accurate and by the queue created by the large number of infected, that could slow down the exams required to be declared recovered. The qualitative fitting can be seen in figure 7.

**Figure 7:**
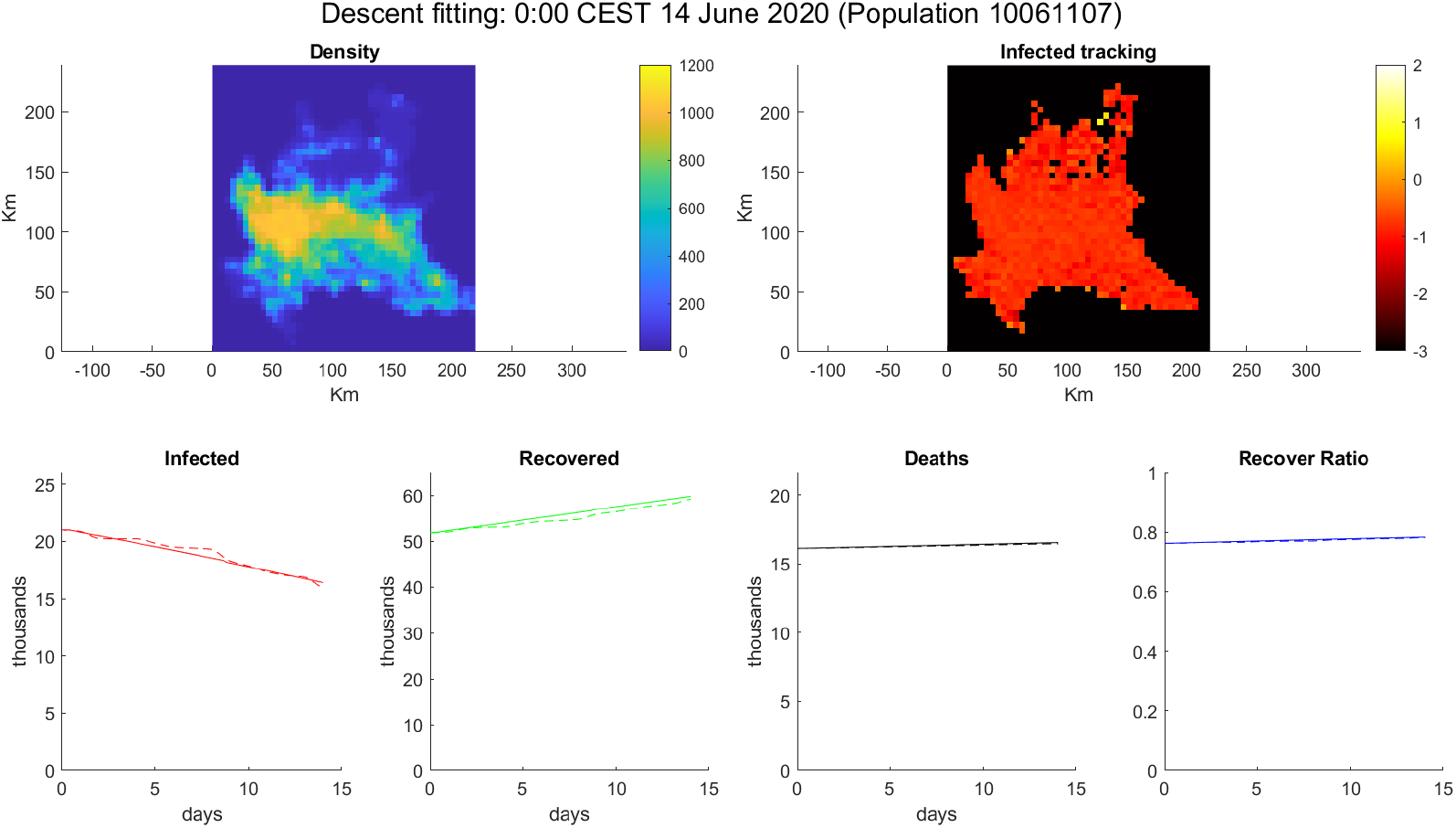
Simulation of descent. (Top-Left) Density of Population. (Top-Right) log_10_ of infected percentage per cell. (Bottom) From left to right: Infected number, Recovered number, Deaths number and Recover ratio 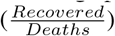. Solid line is model simulation, dotted line is extracted data by [18] for Lombardy.

### 3.5 The vaccine

In the previous scenario of descent has been tested the impact of a vaccination of the 70% of population, near to the 62% suggested in [29]. The agents-based models are perfect to test strategies on the individuals. The result of the simulation is a strong decreasing of infected trend, that is something unexpected in a simulation of 14 days. The results are shown in figure 8.

**Figure 8:**
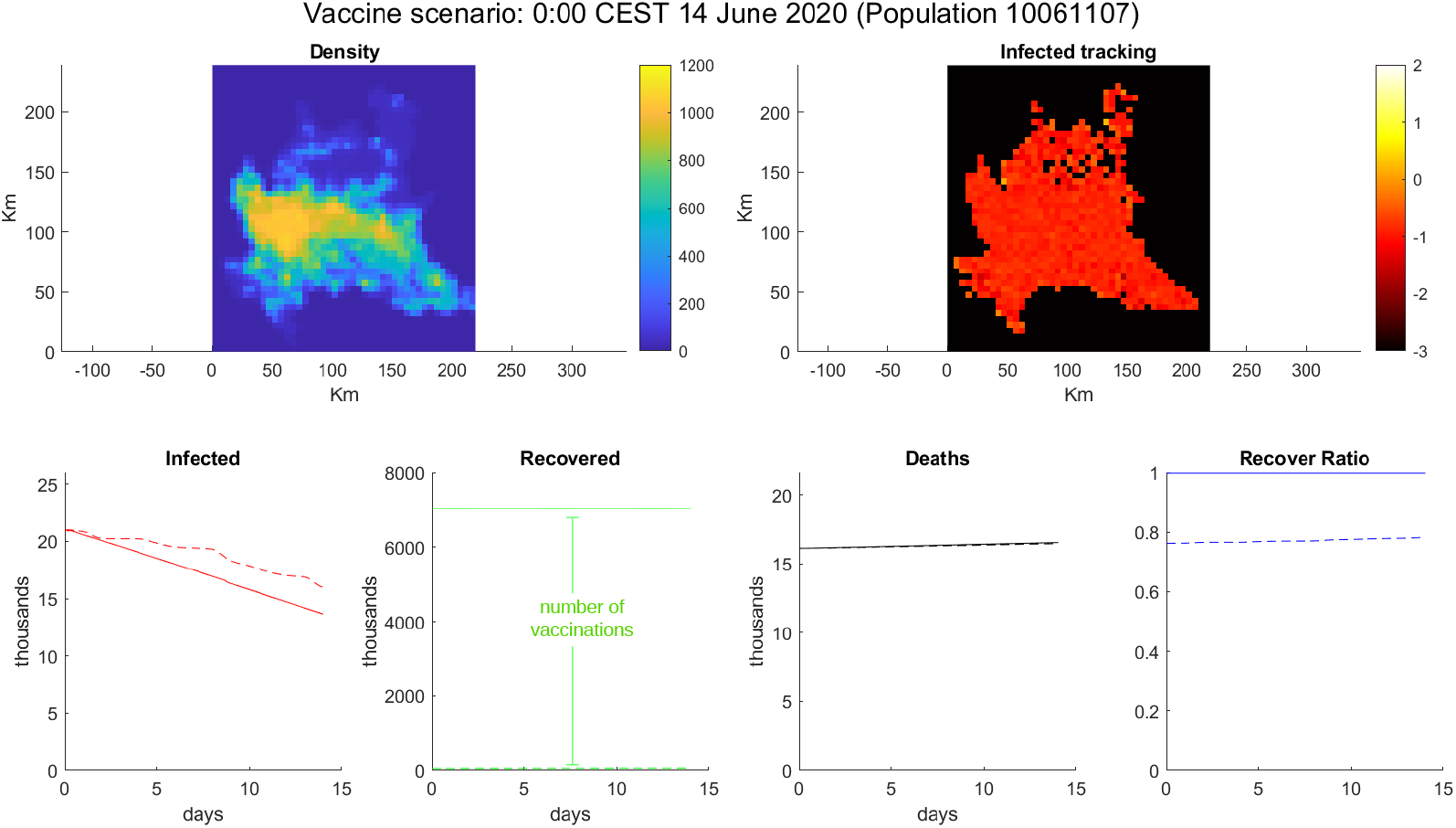
Simulation of vaccination. (Top-Left) Density of Population. (Top-Right) log_10_ of infected percentage per cell. (Bottom) From left to right: Infected number, Recovered number, Deaths number and Recover ratio 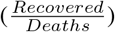. Solid line is model simulation, dotted line is extracted data by [18] for Lombardy.

## 4 Conclusions

This model shows the importance of our simple action in an epidemic setting. Indeed the virus behavior is just an emergent behavior of our habits [10]. However this contribution open a new way in social network analysis, where the graph theoretical approach is substituted by agents. It also open the way to more realistic epidemic models, where the hypothetical scenarios can be tested directly on the agents, without any ODE mediation.

## Data Availability

All the data is open access and cited in the paper. Moreover data can be found at the adress https://github.com/mrjacob241/CTS-Ext, as the source code of the project.

## 5 Acknowledgement

The author received no financial support for the research of this article and all the simulations have been performed on the personal computer of the author. For these reasons doesn’t exist any conflict of interest in publishing the source code of the project in [13]. However author thanks the department of Mathematics and Informatics of the University of Palermo and the Institute of Biophysics of National Research Council (CNR) for funding author’s PhD and for being a constant guide for his growth.

